# P-tau217 and other blood biomarkers of dementia: variation with time of day

**DOI:** 10.1101/2023.12.11.23299805

**Authors:** Ciro della Monica, Victoria Revell, Giuseppe Atzori, Rhiannon Laban, Simon S. Skene, Amanda Heslegrave, Hana Hassanin, Ramin Nilforooshan, Henrik Zetterberg, Derk-Jan Dijk

**Author notes:** Corresponding Author: Derk-Jan Dijk, Surrey Sleep Research Centre, University of Surrey, Guildford, Surrey, GU2 7XP, UK.

## Abstract

Plasma biomarkers of dementia, including phosphorylated tau (p-tau217), offer promise as tools for diagnosis, stratification for clinical trials, monitoring disease progression, and assessing the success of interventions in those living with Alzheimer’s disease. However, currently, it is unknown whether these dementia biomarker levels vary with time of day, which could have implications for their clinical value. In two protocols, we studied 38 participants (70.8 ± 7.6 years; mean ± SD) in a 27-hour laboratory protocol with either two samples taken 12 hours apart or 3-hourly blood sampling for 24 hours in the presence of a sleep-wake cycle. The study population comprised people living with mild Alzheimer’s disease (PLWA, n = 8), partners/caregivers of PLWA (n = 6) and cognitively intact older adults (n = 24). Single molecule array technology was used to measure phosphorylated tau (p-tau217) (ALZpath), amyloid-beta 40 (Aβ40), amyloid-beta 42 (Aβ42), glial fibrillary acidic protein (GFAP), and neurofilament light (NfL) (Neuro 4-Plex E). Analysis with a linear mixed model (SAS, PROC MIXED) revealed a significant effect of time of day for p-tau217, Aβ40, Aβ42, and NfL, and a significant effect of participant group for p-tau217. For p-tau217, lowest levels were observed in the morning upon waking and highest values in the afternoon/early evening. The magnitude of the diurnal variation for p-tau217 was similar to the reported increase in p-tau217 over one year in amyloid-β-positive mild cognitively impaired people. Currently, the factors driving this diurnal variation are unknown and could be related to sleep, circadian mechanisms, activity, posture, or meals. Overall, this work implies that time of day of sample collection may be relevant in the implementation and interpretation of plasma biomarkers in dementia research and care.

## Introduc.on

Alzheimer’s disease (AD) is the most prevalent form of dementia, accounting for up to 80% of all cases, and hallmarks of the disease include amyloid plaques and hyperphosphorylated tau tangles in the brain [1, 2]. There is no standard approach to diagnose AD and disease presence cannot be determined by a single test but is rather a multi-faceted and multi- disciplinary approach involving taking medical history, cognitive tests, amyloid-PET scans, and sometimes cerebrospinal fluid (CSF) samples for measurement of beta-amyloid or tau [2]. However, imaging and CSF tests may not always be possible due to cost, availability of equipment, and the invasiveness of procedures which may not be well tolerated [3]. Nevertheless, the ability to confirm amyloid-beta pathology in the brain will become increasingly important with the advance of disease-modifying therapies (DMTs) targeting amyloid beta e.g., aducanumab [4], lecanemab [5] and donanemab [6], as one of the requirements for prescribing these DMTs is confirmation of brain amyloid burden [7]. Thus, there is a need for acceptable, scalable, and accurate diagnostic approaches to determine disease presence, severity, and response to any treatment.

Plasma biomarkers offer an opportunity as a cost- and pme- effective tool that is minimally invasive for screening and diagnosis, stratification, monitoring disease progression, and assessing treatment response. Biomarkers that have been proposed include amyloid-beta (Aβ40, Aβ42, and their ratio), phosphorylated tau (p-tau181, p- tau217), glial fibrillary acidic protein (GFAP), and neurofilament light (NfL) (reviewed in [1]). The sensitivity and specificity of these biomarkers is an area of active research. In particular, p-tau217 has been demonstrated to be a valuable biomarker for predicting cognitive decline and monitoring treatment efficacy in response to DMT [8, 9].

Although plasma biomarkers, and particularly p-tau217, show great promise as clinical tools very liyle is known about non-disease related factors that may influence the concentrations of these biomarkers in blood. Biomarker levels may vary between individuals due to demographic or comorbid factors (inter-individual variation), but they may also vary within an individual due to behaviour or biological processes (intra-individual variation). Factors of interest include demographic variables such as age and sex, but also behavioural factors such as activity, posture, and eating and drinking. One factor of particular interest is pme-of-day since many physiological variables in blood display 24-h rhythmicity. However, to date, the impact of time-of-day has not been taken into consideration for the implementation of plasma biomarkers for dementia. The importance of pme of day for diagnostic samples has already been demonstrated in other clinical conditions. For example, for people living with severe asthma, sputum samples from morning clinic have significantly higher levels of eosinophils than samples from azernoon clinics [10] which may impact on clinical decision making.

Here, in a controlled laboratory se{ng, we explored whether plasma levels of biomarkers of dementia-related brain changes over the course of a 24-hour day (that includes a sleep/wake cycle and meals) in a heterogenous group of participants consisting of people living with mild Alzheimer’s disease (PLWA), their caregivers, and cognitively intact older adults.

## Materials and methods

### Participants

#### Demographics

Data were collected from participants who were enrolled in one of two studies: 1) in cognitively intact older adults, 2) in PLWA, their study partner, and cognitively intact older adults. Eligibility was assessed using pre-defined inclusion/exclusion criteria for each of the three study groups. PLWA had to be 50 – 85 years old with a confirmed diagnosis of prodromal or mild AD, have an SMMSE (Standardised Mini Mental State Examination [11]) score ≥ 23, be living in the community and be on a stable dose of any medication for dementia for at least three months. The diagnosis of prodromal or mild AD was based on clinical history, cognitive tests, and CT/MRI imaging. PLWA could participate in the study by themselves, or they could have a ‘study partner’ who must have known them for at least six months and could be their carer or a family member or friend. Study partners were ≥ 18 years and had to have an SMMSE score ≥ 27. Cognitively intact older adults had to be aged 50 – 85 years, have an SMMSE score ≥ 27 (Study 2), and any comorbidities and concomitant medications must have been stable for the past three months. Cognitively intact adults were recruited via the Surrey Clinical Research Facility database. Potentially eligible PLWA and their study partners were identified via Surrey and Borders Partnership NHS Foundation Trust (SABP) memory services and were approached by one of the SABP team initially by telephone to discuss the study before being provided with the participant information sheet.

#### Ethics

Study One (cognitively intact older adults) received a favourable opinion from the University of Surrey Ethics Committee, and Study Two (PLWA, caregivers of PLWA, and cognitively intact older adults) received a favourable opinion from an NHS ethics committee (22/LO/0694). Study Two is ongoing and is registered as a clinical study on the ISRCTN (International Standard Randomised Controlled Trial Number) registry (ISRCTN10509121). The protocols were conducted in accordance with the Declaration of Helsinki and guided by the principles of Good Clinical Practice. All personal data was handled in accordance with the general data protection regulations (GDPR) and the UK Data Protection Act 2018.

Written informed consent was obtained from participants prior to any study procedures being performed. Participants were compensated for their time and inconvenience.

### Procedures & Measures

The full study protocols have been reported in detail elsewhere [12, 13]. Briefly, following a screening visit to assess eligibility, participants were monitored for up to 14 days at-home using a variety of technologies to assess their sleep-wake patterns, environment, and cognitive function. They then attended the UK-DRI Clinical Research Facility at the University of Surrey for a 27-hour residential session which included a full clinical polysomnography (PSG) recording during an extended 10-hour period in bed. PSG was recorded using the Somnomedics SomnoHD system with Domino software (v 3.0.0.6; sampled at 256 Hz; SOMNOmedics GmbHTM, Germany) with an American Academy of Sleep Medicine (AASM) standard adult montage.

During the residential session, participants remained in environmentally controlled bedroom environments with en-suite facilities. For PLWA and their study partners, the aim was to recreate their sleeping situation at home so they could either share a room in a double occupancy suite or could be in adjacent rooms with an interconnecting door. During the afternoon/evening/morning hours participants were free to pursue their own activities around scheduled procedures including sample collection, meals, questionnaire completion, and having PSG equipment attached. In Study One, dinner was scheduled ∼5 hours before habitual bedtime (at approximately 18:30 h) and breakfast ∼2 hours after habitual waketime (at approximately 09:30 h). In Study Two, lunch was ∼9.5 hours and dinner was ∼4 hours before habitual bedtime; breakfast was ∼1.5 hours after habitual waketime.

In Study One, all participants had two blood samples drawn 12 hours apart via venepuncture at 19:46 ± 00:33 h and 07:53± 00:35 h (mean ± SD). The evening sample was 2.88 ± 0.80 hours before Lights Off, and the morning sample was 0.68 ± 0.92 hours after Lights On. In Study Two, participants had an indwelling cannula sited and blood samples were drawn at three-hourly intervals relative to their habitual bedtime. Sampling began nine hours before habitual bedtime and continued until 15 hours after habitual bedtime.

Blood samples were collected into potassium EDTA tubes, were centrifuged immediately upon collection, and the plasma fraction was separated and stored at -80^0^C. Samples were shipped to the UK DRI Biomarker Factory, UCL, London where they were analysed using Simoa HD-X technology. The following biomarkers were measured in both studies using the Neuro 4-PlexE assay kit (Quanterix, Billerica MA): amyloid-beta 40 (Ab40), amyloid-beta 42 (Ab42), glial fibrillary acidic protein (GFAP), neurofilament light (NfL). Tau phosphorylated at threonine 217 (p-tau217) was measured using the ALZpath Simoa assay and measured in Study two only (ALZpath, Carlsbad, CA). Samples were measured in singlicate, and four internal controls made of pooled plasma were used to monitor any intra-and inter-plate variation. All coefficients of variation for internal controls were below 10%.

## Data analysis

The data were analysed for all participants combined and separately for each group (PLWA, Study Partner, Cognitively intact) and, for each of the five biomarkers as well as the Aβ42/Aβ40 ratio. For each biomarker, the mean values at each timepoint were computed and also intraclass correlations (ICCs) were calculated using R Statistical Software (v4.2.2; R Core Team 2022). A PROC MIXED linear model was run in SAS (v9.4, SAS Institute Inc), and LS (least squares) means generated, using the Study Two datasets (nine timepoints) or the combined Study One and Two datasets (two timepoints) to assess the impact of time-of-day, group, and any interaction. For the two timepoint comparison, the timepoints used for the Study Two datasets were three hours before and nine hours after habitual bedtime; p- tau217 was only from Study Two but all other biomarkers were combined Study One and Two datasets. An extended PROC MIXED linear model was run for both the nine and two timepoint datasets where in addition to time and group, the following five factors were included as covariates: age, sex, BMI, PSQI, and AHI.

## Results

Here we report data from 38 participants. The cognitively intact adults included 17 from study one (72.0±4.5 years; 11M: 6F; 28.9±1.3 SMMSE; 26.4±4.7 kg/m^2^ BMI) and seven from study two (67.0±6.2 years; 4M: 3F; 28.9±1.1 SMMSE; 26.4±3.8 kg/m^2^ BMI). We enrolled eight PLWA (74.8±4.4 years; 4M: 4F; 27.0±1.8 SMMSE; 29.7±7.6 kg/m^2^ BMI), and six partners of PLWA (66.7±14.8 years; 3M: 3F; 28.8±1.0 SMMSE; 28.9±4.2 kg/m^2^ BMI). Comorbidities included hypertension, Type-2 diabetes, arthritis, hyperthyroidism, and asthma [12, 13]

For both studies combined, 90% of scheduled samples were obtained. The plasma levels for each biomarker at each timepoint (mean ± SD) are given in Table 1 for all participants combined as well as separately for each group for the nine timepoint comparison (Study Two). The ICCs for all participants combined ranged between 0.84 and 0.97 for the different biomarkers and the values were similar across groups. Table 2 provides a similar comparison for the two timepoint comparison, and here the ICCs range from 0.76 to 0.93 for all participants combined. These ICC values imply that the between-participant variation is greater than the within-participant variation and that this is similar across groups.

**Table 1.**
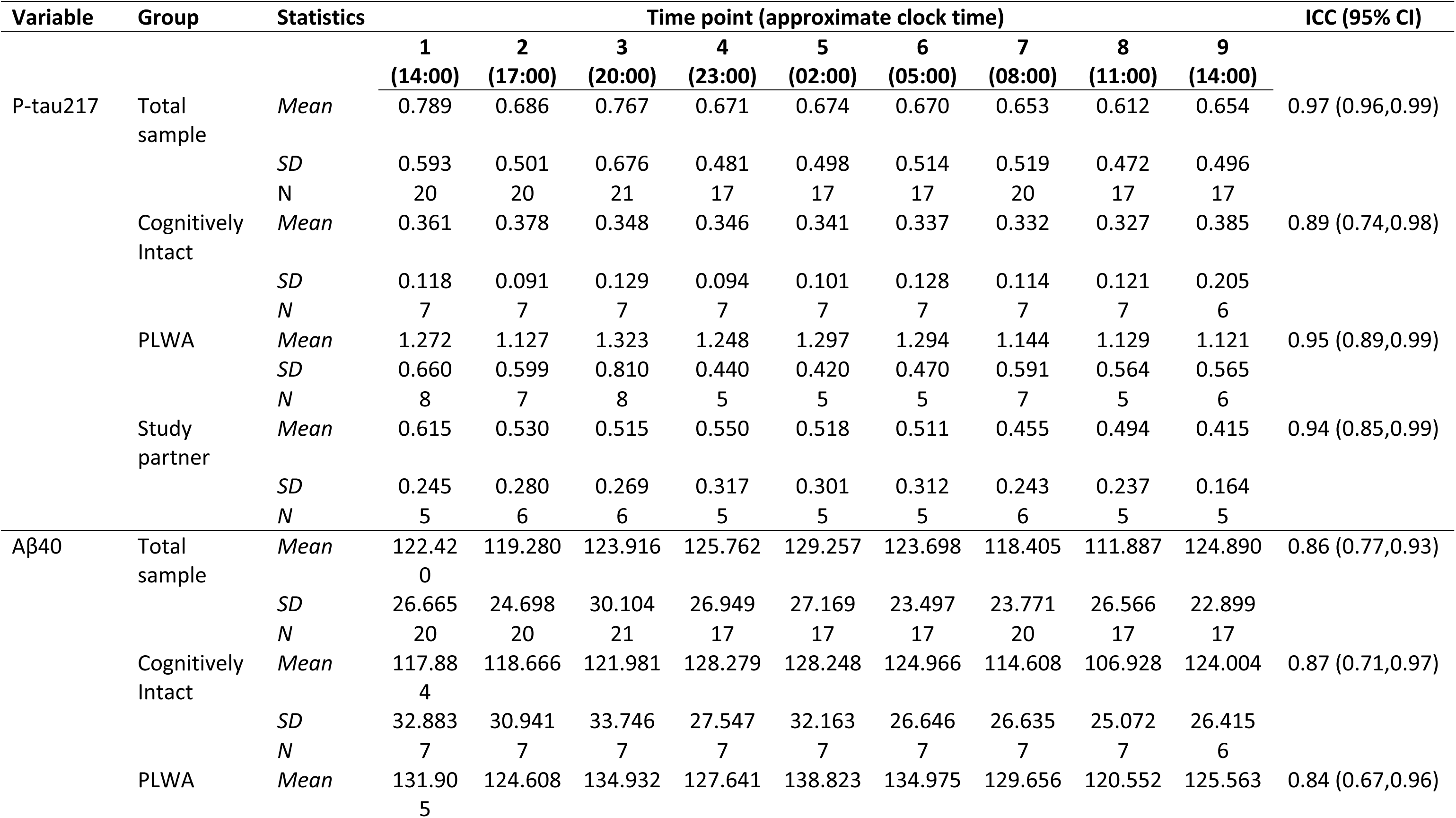
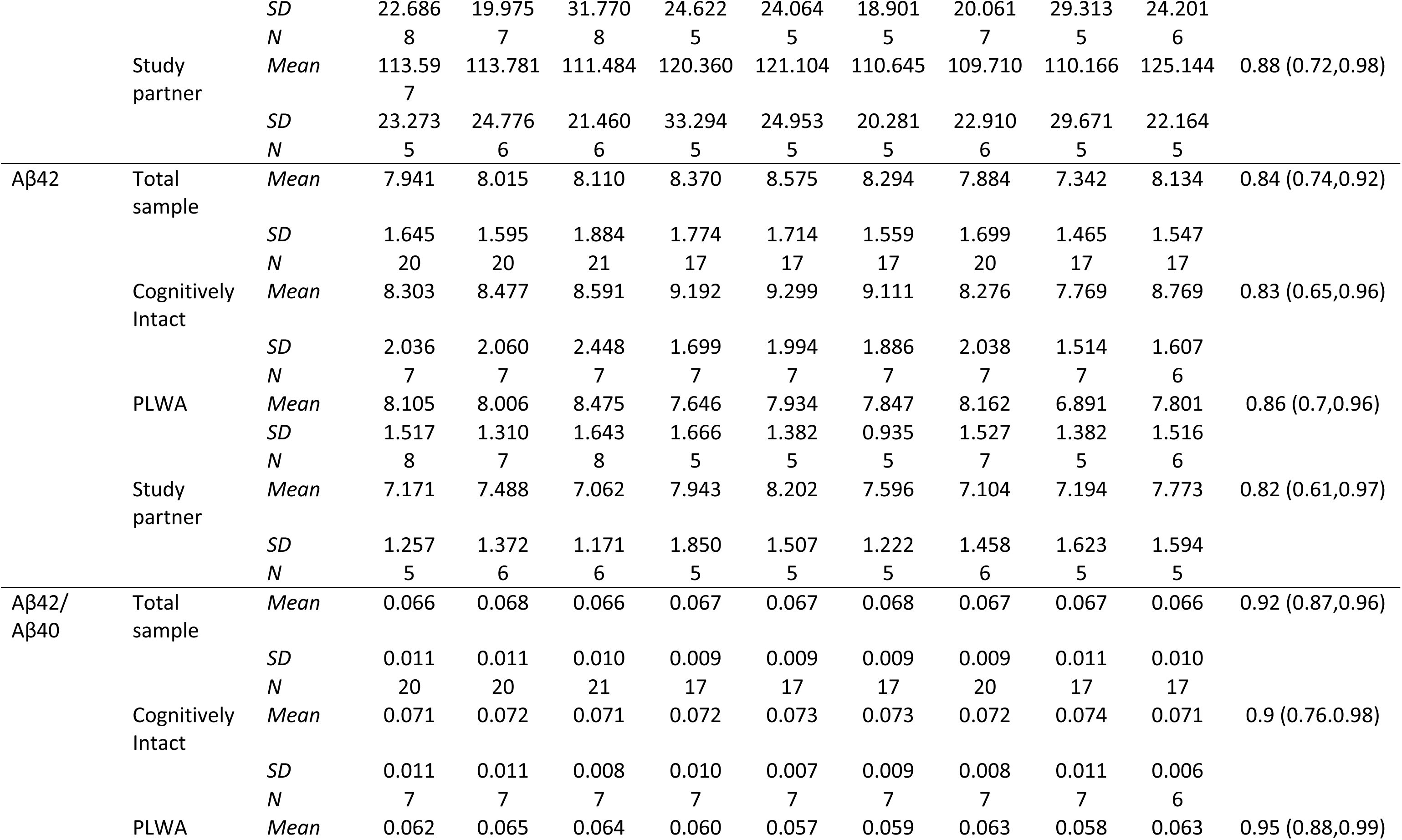
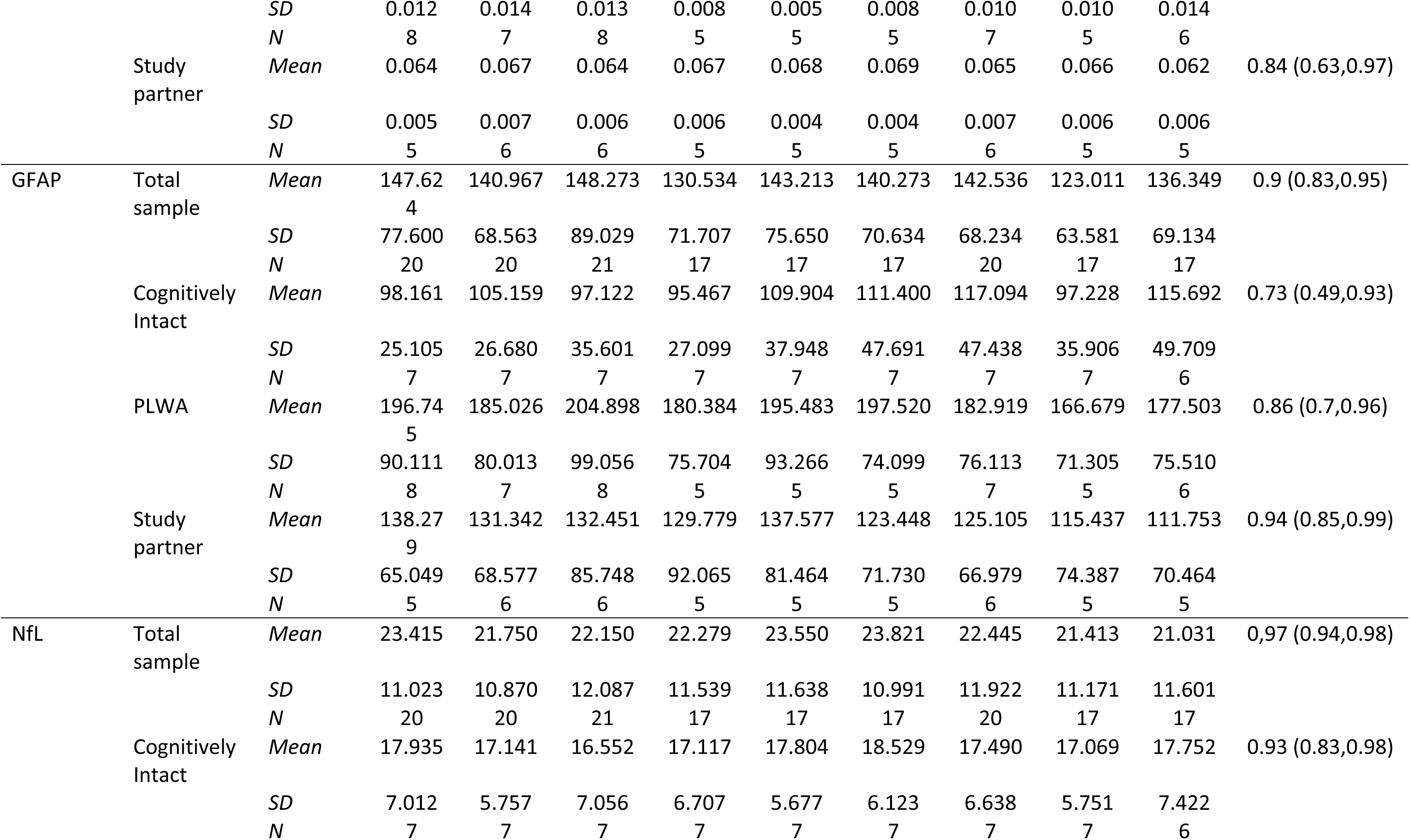
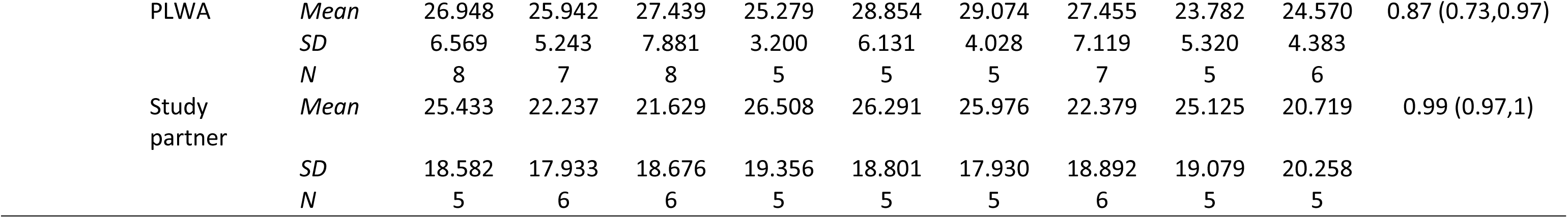
Plasma biomarker levels (pg/mL, mean ± SD) across a 24-hour period.

**Table 2.**
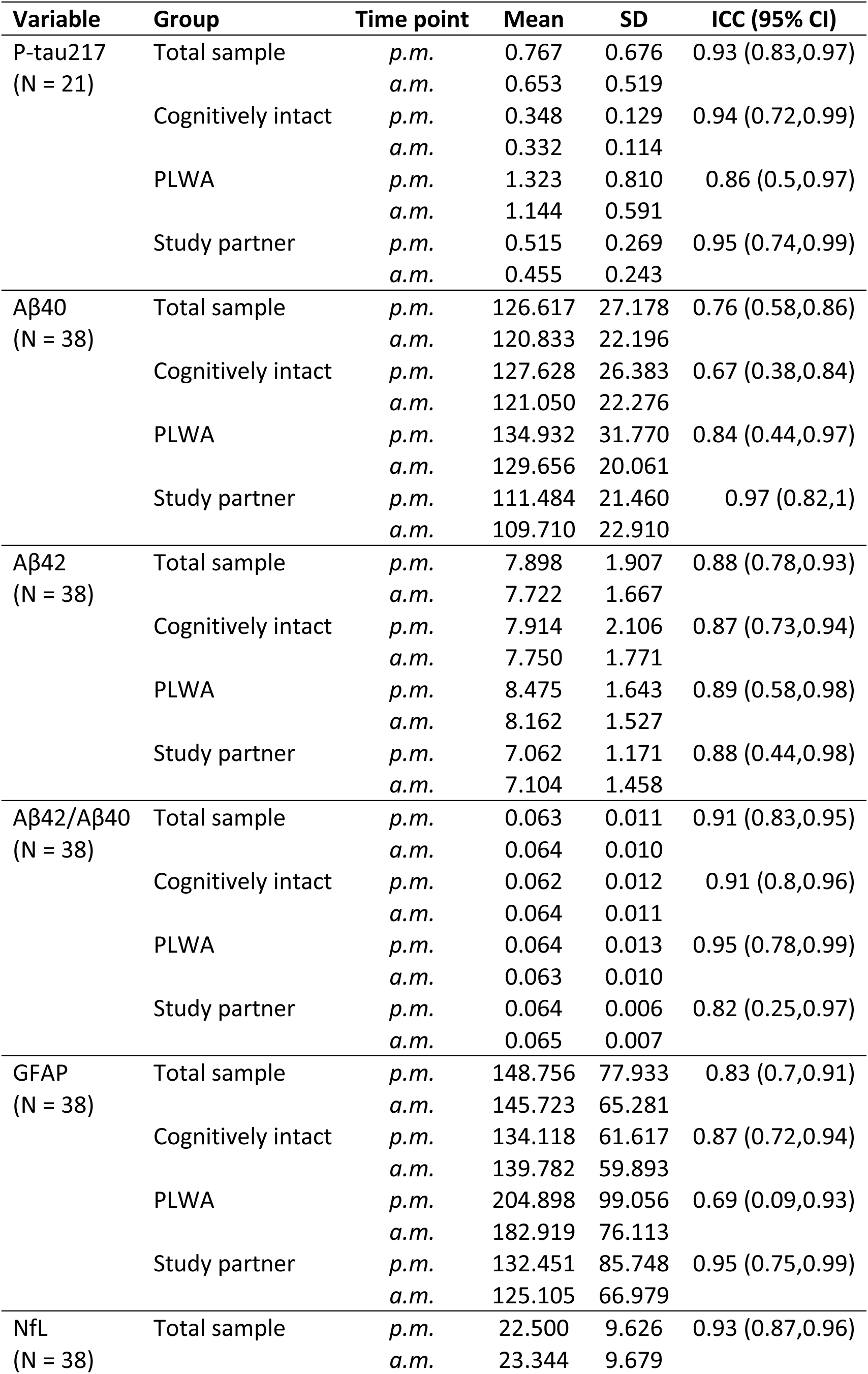
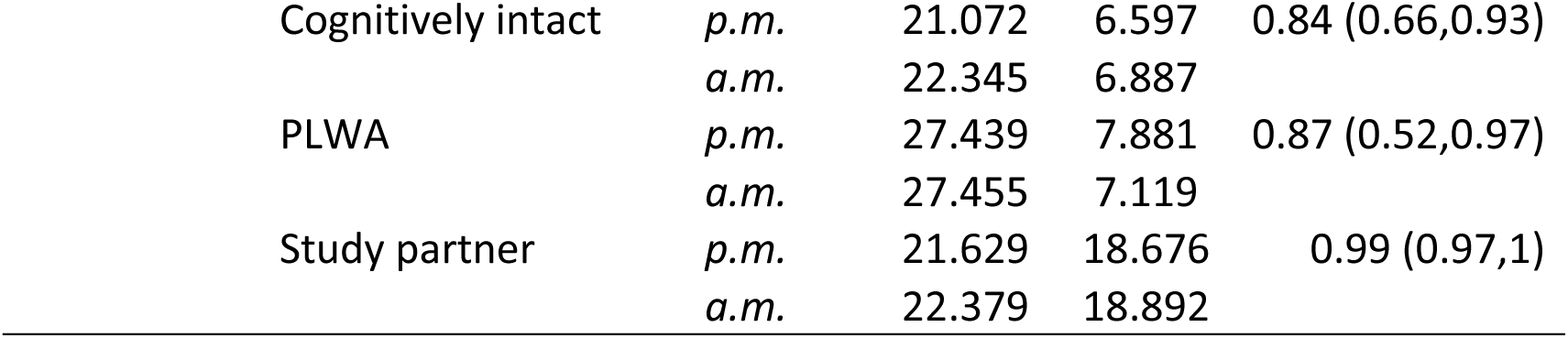
Plasma biomarker levels (pg/mL, mean ± SD): evening vs morning.

| Variable | Group | Time point | Mean | SD | ICC (95% CI) |
| --- | --- | --- | --- | --- | --- |
| P-tau217<br>(N = 21) | Total sample | *p.m.* | 0.767 | 0.676 | 0.93 (0.83,0.97) |
|  |  | *a.m.* | 0.653 | 0.519 |  |
|  | Cognitively intact | *p.m.* | 0.348 | 0.129 | 0.94 (0.72,0.99) |
|  |  | *a.m.* | 0.332 | 0.114 |  |
|  | PLWA | *p.m.* | 1.323 | 0.810 | 0.86 (0.5,0.97) |
|  |  | *a.m.* | 1.144 | 0.591 |  |
|  | Study partner | *p.m.* | 0.515 | 0.269 | 0.95 (0.74,0.99) |
|  |  | *a.m.* | 0.455 | 0.243 |  |
| Aβ40<br>(N = 38) | Total sample | *p.m.* | 126.617 | 27.178 | 0.76 (0.58,0.86) |
|  |  | *a.m.* | 120.833 | 22.196 |  |
|  | Cognitively intact | *p.m.* | 127.628 | 26.383 | 0.67 (0.38,0.84) |
|  |  | *a.m.* | 121.050 | 22.276 |  |
|  | PLWA | *p.m.* | 134.932 | 31.770 | 0.84 (0.44,0.97) |
|  |  | *a.m.* | 129.656 | 20.061 |  |
|  | Study partner | *p.m.* | 111.484 | 21.460 | 0.97 (0.82,1) |
|  |  | *a.m.* | 109.710 | 22.910 |  |
| Aβ42<br>(N = 38) | Total sample | *p.m.* | 7.898 | 1.907 | 0.88 (0.78,0.93) |
|  |  | *a.m.* | 7.722 | 1.667 |  |
|  | Cognitively intact | *p.m.* | 7.914 | 2.106 | 0.87 (0.73,0.94) |
|  |  | *a.m.* | 7.750 | 1.771 |  |
|  | PLWA | *p.m.* | 8.475 | 1.643 | 0.89 (0.58,0.98) |
|  |  | *a.m.* | 8.162 | 1.527 |  |
|  | Study partner | *p.m.* | 7.062 | 1.171 | 0.88 (0.44,0.98) |
|  |  | *a.m.* | 7.104 | 1.458 |  |
| Aβ42/Aβ40<br>(N = 38) | Total sample | *p.m.* | 0.063 | 0.011 | 0.91 (0.83,0.95) |
|  |  | *a.m.* | 0.064 | 0.010 |  |
|  | Cognitively intact | *p.m.* | 0.062 | 0.012 | 0.91 (0.8,0.96) |
|  |  | *a.m.* | 0.064 | 0.011 |  |
|  | PLWA | *p.m.* | 0.064 | 0.013 | 0.95 (0.78,0.99) |
|  |  | *a.m.* | 0.063 | 0.010 |  |
|  | Study partner | *p.m.* | 0.064 | 0.006 | 0.82 (0.25,0.97) |
|  |  | *a.m.* | 0.065 | 0.007 |  |
| GFAP<br>(N = 38) | Total sample | *p.m.* | 148.756 | 77.933 | 0.83 (0.7,0.91) |
|  |  | *a.m.* | 145.723 | 65.281 |  |
|  | Cognitively intact | *p.m.* | 134.118 | 61.617 | 0.87 (0.72,0.94) |
|  |  | *a.m.* | 139.782 | 59.893 |  |
|  | PLWA | *p.m.* | 204.898 | 99.056 | 0.69 (0.09,0.93) |
|  |  | *a.m.* | 182.919 | 76.113 |  |
|  | Study partner | *p.m.* | 132.451 | 85.748 | 0.95 (0.75,0.99) |
|  |  | *a.m.* | 125.105 | 66.979 |  |
| NfL<br>(N = 38) | Total sample | *p.m.* | 22.500 | 9.626 | 0.93 (0.87,0.96) |
|  |  | *a.m.* | 23.344 | 9.679 |  |

|  |  |  |  |  |
| --- | --- | --- | --- | --- |
| Cognitively intact | *p.m.* | 21.072 | 6.597 | 0.84 (0.66,0.93) |
|  | *a.m.* | 22.345 | 6.887 |  |
| PLWA | *p.m.* | 27.439 | 7.881 | 0.87 (0.52,0.97) |
|  | *a.m.* | 27.455 | 7.119 |  |
| Study partner | *p.m.* | 21.629 | 18.676 | 0.99 (0.97,1) |
|  | *a.m.* | 22.379 | 18.892 |  |

Two PROC MIXED models using two datasets were run as well as an extended model as described in the Data Analysis section. The model using the nine timepoint comparison showed that there was a significant main effect of time for all biomarkers (p < 0.01) except GFAP (p = 0.065) (Table 3). Figure 1 shows plasma biomarker levels (LS-means) for all participants across 24-hours. For plasma p-tau217 (LS-means) lowest values were observed in the morning, shortly after wake time, after which levels rose to highest values in the afternoon and evening. Thus, p-tau217 concentrations in the first two samples after wake time were significantly lower (p < 0.0001) compared to the evening (3 hours before habitual bedtime) sample. For Aβ40 and Aβ42, higher levels were observed in the afternoon/evening hours with peak levels during the sleep episode and lowest levels in the morning hours. NfL levels peaked overnight with similar levels in the evening and morning hours. The magnitude of the diurnal variation (change in LS-means expressed as a percentage from the overall mean) was: 14.0% (Aβ40), 15.3% (Aβ42), 4.6% (Aβ42/ Aβ40), 10.6% (NfL), 17.0% (GFAP), 15.8% (p-tau217).

**Figure 1.**
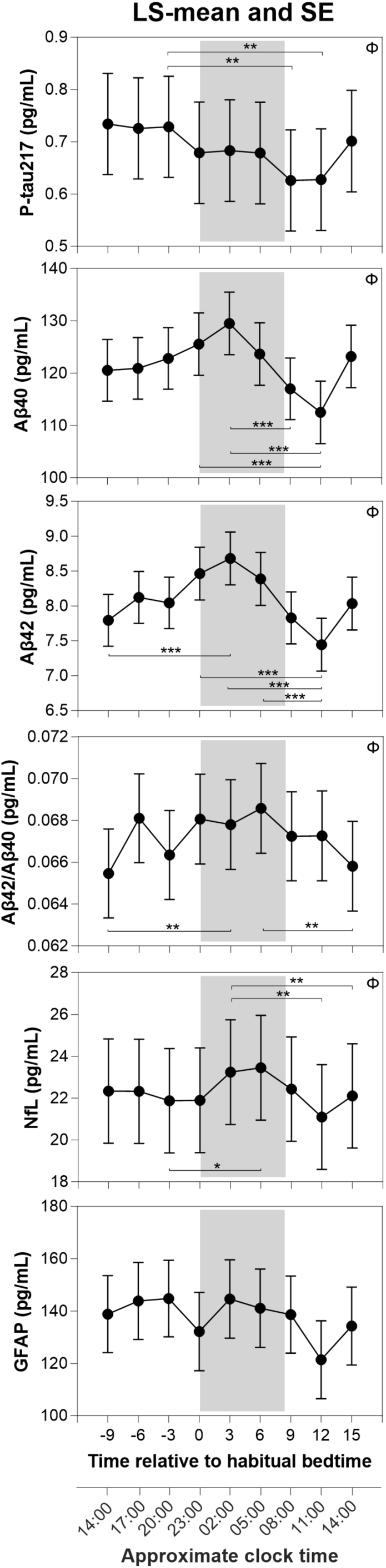
Levels of plasma biomarkers (LS-means +/- SE) across a 24-hour period: p-tau217, Aβ40, Aβ42, Aβ42/Aβ40, NfL, GFAP. The grey shading indicates the habitual sleep episode. <Ι indicates an overall significant effect of time. Asterisk symbol indicates a significant difference in levels between the indicated timepoints; *** = p<0.0001, ** = p<0.01, * = p<0.05.

**Table 3.**
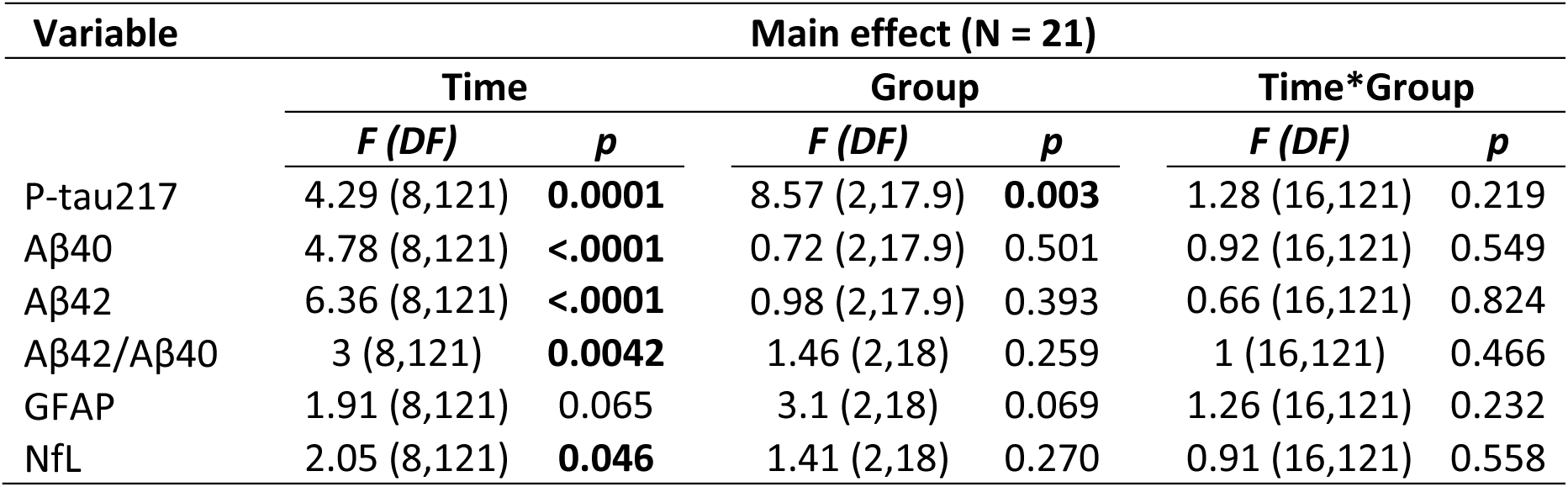
Summary of PROC MIXED analysis for plasma biomarkers over nine timepoints.

A significant effect of group was observed for p-tau217 (p = 0.003) (Figure 2) with highest levels observed in PLWA, and the effect of group approached significance for GFAP (p = 0.069). For none of the biomarkers a significant group by time interaction was observed. For p-tau217, the magnitude of the diurnal variation in PLWA estimated from the LS-means (0.233 ± 0.044, LS-mean ± SE) was 27% of the difference between the mean values for cognitively intact adults (0.349 ± 0.164, LS-mean ± SE) and PLWA (1.215 ± 0.153, LS- mean ± SE).

**Figure 2.**
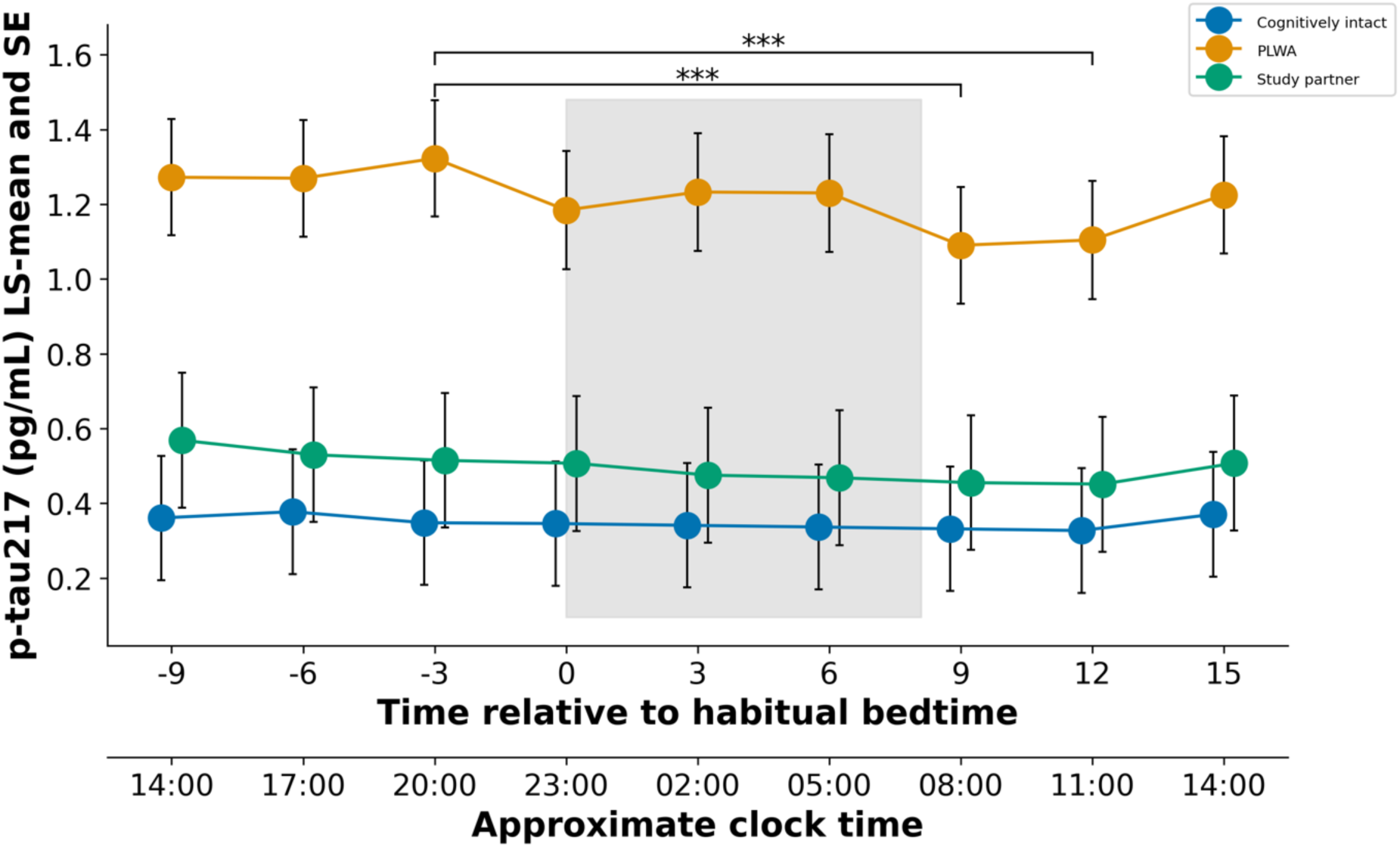
Levels of plasma p-tau217 (LS-means +/- SE) across a 24-hour period in PLWA, their Study Partners, and Cognitively Intact older adults. Blue symbols represent cognitively intact older adults, green symbols represent Study Partners, and orange symbols represent PLWA. The grey shading indicates the habitual sleep episode. *** indicates a significant (p < 0.0001) difference in levels between the indicated timepoints in PLWA. The data for the Study Partners and the Cognitively Intact older adults are displaced by 15 minutes so that the variance indicators of the various groups are visible.

To further establish the effects of time we compared evening to morning samples. In this comparison data were available for 38 participants for all biomarkers except p-tau217 (n = 21). PROC MIXED analysis on the two timepoints only (Table 4) revealed significant effects of time, group, and group by time interaction for p-tau217 only.

**Table 4.** Summary of PROC MIXED analysis for plasma biomarkers over two timepoints.

| Variable | N | Main effect |  |  |  |  |  |
| --- | --- | --- | --- | --- | --- | --- | --- |
|  |  | Time |  | Group |  | Time*Group |  |
|  |  | *F* (DF) | *p* | *F* (DF) | *P* | *F* (DF) | *p* |
| P-tau217 | 21 | 7.77 (1,17) | **0.013** | 7.35 (2,17.9) | **0.005** | 3.66 (2,17) | **0.048** |
| Aβ40 | 38 | 3.86 (1,32.6) | 0.058 | 1.19 (2,34.2) | 0.315 | 0.42 (2,32.6) | 0.662 |
| Aβ42 | 38 | 1.89 (1,32.9) | 0.179 | 0.73 (2,34.7) | 0.488 | 0.54 (1,32.9) | 0.5869 |
| Aβ42/Aβ40 | 38 | 1.38 (1,33.1) | 0.248 | 0.09 (2,35) | 0.914 | 0.18 (2,331.1) | 0.839 |
| GFAP | 38 | 2.9 (1,33) | 0.098 | 1.9 (2,34.8) | 0.165 | 2.87 (2,33) | 0.071 |
| NfL | 38 | 0.27 (1,33.2) | 0.606 | 1.03 (2,35.1) | 0.367 | 0.72 (2,33.2) | 0.495 |

When the extended PROC MIXED model was run for the nine timepoint dataset, the effects of time remained significant for p-tau217, Aβ40, Aβ42, Aβ42/ Aβ40, and NfL. For p- tau217 the effect of group remained significant and a significant interaction between time and group emerged. A significant effect of age was observed for GFAP (p = 0.036). No significant effects of sex, BMI, PSQI or AHI were observed for any of the biomarkers (Supplementary Table 1). For the two timepoint dataset (Supplementary Table 2), a similar significant effect of age was observed for GFAP (p = 0.006) and for p-tau217 the significant effects of time, group, and group*time remained.

## Discussion

Here, we show that levels of commonly used plasma biomarkers in dementia research including p-tau217, Aβ40, Aβ42, Aβ42/Aβ40, and NfL vary with time-of-day. This significant variation with time-of-day was observed despite the rather large ICC values (range 0.76 – 0.97), which indicate that the between participant variation is greater than within participant variation. The ICCs reported here are in line with previous studies which investigated the longitudinal reliability of plasma biomarkers and observed ICC values between 0.66 and 0.78 [14]. Our observed impact of age on GFAP levels is in line with previous observations in people living with Parkinson’s disease where GFAP was shown to correlate with both age and MMSE [15].

We observed significant time-of-day variation for p-tau217, NfL, Aβ40, Aβ42, and Aβ42/Aβ40 with the effect approaching significance for GFAP, with the magnitude of diurnal variation ranging from 4.6 – 15.8% for the significant effects. Previous work has demonstrated that cerebrospinal (CSF) levels of amyloid-beta fluctuate with time of day [16–18]. The observed diurnal fluctuations for Aβ40 and Aβ42 were 2.6% and 0.4%, respectively, for amyloid-positive participants, with the highest values in the early afternoon and lowest values upon waking [17]. This compares to a 14.0% for Aβ40 and 15.3% for Aβ42 diurnal variation in plasma observed in the current study.

The shape of the diurnal variation varied across the biomarkers. For p-tau217 highest values were observed before bedtime and lowest values upon awakening. For Aβ40, and Aβ42, we observed highest values during the nocturnal sleep period and lowest values upon waking. A previous study of Aβ40, and Aβ42 in CSF showed levels were lower in the morning with highest values were in the afternoon [17]. For NfL, highest values were also observed during the sleep period with lowest values in mid-morning and relatively stable levels in the afternoon/evening and morning. Larger sample sizes are needed to further determine the precise shape of this diurnal variation and differences therein across the biomarkers.

The factors underlying the observed diurnal variation remain to be identified. They could be related to circadian modulation of production, phosphorylation, clearance from the brain or could be a response to behavioural changes/processes across the 24-hour day including sleep, meals, or posture. In the latter case, simple behavioural constraints could remove the variance and samples could be taken at any time, whereas in the former case, sample should be taken within particular time windows or values should be corrected for time of day. The observed differences in the shape and timing of the diurnal variation across the biomarkers make it unlikely that one common mechanism, such as changes in blood volume, or circadian or sleep mediated clearance from the brain into the circulation drives all of this diurnal variation.

Although the time-of-day effects we observed may appear small, when they are placed in the context of disease or treatment monitoring, they become of clinical interest. For example, plasma p-tau217 has recently become a biomarker of interest in AD research due to its sensitivity for discriminating for AD, its ability to predict cognitive decline, and its capacity to track response to DMT [8, 9, 19]. Of particular interest is a study in cohorts of Aβ positive individuals (n = 171) who were cognitively unimpaired [8]. In this study cognition was assessed using the mini-mental state examination (MMSE) and the modified preclinical Alzheimer Cognitive Composite (mPACC) over a median of six years. Plasma p-tau217 was shown to be the strongest biomarker for predicting cognitive decline and also conversion to AD [8]. Of particular relevance to our findings is that longitudinal monitoring in those with Aβ-positive prodromal AD showed an increase in p-tau217 of 14.7% per year [20]. This is very similar to the magnitude of the diurnal variation (15.8%) observed in the current study. This change is also meaningful when we consider that in the TRAILBLAZER-ALZ clinical trial following treatment with donanemab for up to 72 weeks, plasma-tau217 levels declined by 23% [9] and GFAP levels decreased by 12%, whereas under placebo both biomarkers increased by 6 and 15%. These percentages are also within the range of the systematic effect of pme of day observed in our study.

Of the plasma biomarkers assessed in the TRAILBLAZER Trial (GFAP, NfL, p-tau217, and Aβ42/Aβ40) only p-tau217 was positively and significantly associated with baseline amyloid plaques and global tau deposition. It is of interest that in our small sample only p- tau217 showed a significant group effect.

For now, our results suggest that time of day matters when considering sampling for plasma biomarkers of dementia for monitoring disease progression or treatment outcome. Samples obtained at an early morning clinic may provide different results to those taken in an afternoon or evening clinic. Time of day should be standardised or at least recorded when samples are collected whether for diagnosis or monitoring their clinical status longitudinally. Recent studies suggest that biomarker concentrations also vary by food intake [21]. For now, we recommend that reference limits for biomarkers related to neurodegenerative dementias are established in samples collected fasting and in the morning, and that samples for dementia diagnostics are collected accordingly.

## Data Availability

All data produced in the present study are available upon reasonable request to the authors.

## Acknowledgements

This work is supported by the UK Dementia Research Institute, Care Research and Technology Centre at Imperial College, London and the University of Surrey, Guildford, United Kingdom which receives its funding from UK DRI Ltd, funded by the UK Medical Research Council, as well as the National Institute for Health and Care Research University College London Hospitals Biomedical Research Centre, and the UK Dementia Research Institute at UCL (UKDRI-1003), Alzheimer’s Society and Alzheimer’s Research UK. We thank Surrey Clinical Research Facility for support with recruitment and residential sessions, in particular Kat Pizzoferro for her oversight of sample collection and processing.

## References

1. Blennow K, Galasko D, Perneczky R, Quevenco FC, van der Flier WM, Akinwonmi A et al. The potential clinical value of plasma biomarkers in Alzheimer’s disease. Alzheimers Dement 2023.

2. 2020 Alzheimer’s disease facts and figures. Alzheimers Dement 2020.

3. Bittner T. Editorial: What Are the Remaining Challenges before Blood-Based Biomarkers for Alzheimer’s Disease Can Be Used in Clinical Practice? J Prev Alzheimers Dis 2022; 9(4): 567-568.

4. Budd Haeberlein S, Aisen PS, Barkhof F, Chalkias S, Chen T, Cohen S et al. Two Randomized Phase 3 Studies of Aducanumab in Early Alzheimer’s Disease. J Prev Alzheimers Dis 2022; 9(2): 197–210.

5. van Dyck CH, Swanson CJ, Aisen P, Bateman RJ, Chen C, Gee M et al. Lecanemab in Early Alzheimer’s Disease. N Engl J Med 2023; 388(1): 9–21.

6. Sims JR, Zimmer JA, Evans CD, Lu M, Ardayfio P, Sparks J et al. Donanemab in Early Symptomatic Alzheimer Disease: The TRAILBLAZER-ALZ 2 Randomized Clinical Trial. JAMA 2023; 330(6): 512–527.

7. Cummings J, Aisen P, Apostolova LG, Atri A, Salloway S, Weiner M. Aducanumab: Appropriate Use Recommendations. J Prev Alzheimers Dis 2021; 8(4): 398–410.

8. Mattsson-Carlgren N, Salvado G, Ashton NJ, Tideman P, Stomrud E, Zetterberg H et al. Prediction of Longitudinal Cognitive Decline in Preclinical Alzheimer Disease Using Plasma Biomarkers. JAMA Neurol 2023; 80(4): 360–369.

9. Pontecorvo MJ, Lu M, Burnham SC, Schade AE, Dage JL, Shcherbinin S et al. Association of Donanemab Treatment With Exploratory Plasma Biomarkers in Early Symptomatic Alzheimer Disease: A Secondary Analysis of the TRAILBLAZER-ALZ Randomized Clinical Trial. JAMA Neurol 2022; 79(12): 1250–1259.

10. Durrington HJ, Gioan-Tavernier GO, Maidstone RJ, Krakowiak K, Loudon ASI, Blaikley JF et al. Time of Day Affects Eosinophil Biomarkers in Asthma: Implications for Diagnosis and Treatment. Am J Respir Crit Care Med 2018; 198(12): 1578–1581.

11. Molloy DW, Standish TI. A guide to the standardized Mini-Mental State Examination. Int Psychogeriatr 1997; 9 **Suppl 1**: 87–94; discussion 143-150.

12. Ravindran GKK, della Monica C, Atzori G, Lambert D, Hassanin H, Revell V et al. Three Contactless Sleep Technologies Compared to Actigraphy and Polysomnography in a Heterogenous Group of Older Men and Women in a Model of Mild Sleep Disturbance: A Sleep Laboratory Study. JMIR mHealth and uHealth 2023; 25/08/2023:46338 - (forthcoming/in press).

13. Ravindran KKG, della Monica C, Atzori G, Lambert D, Hassanin H, Revell V et al. Contactless and Longitudinal Monitoring of Nocturnal Sleep and Daytime Naps in Older Men and Women: A Digital Health Technology Evaluation Study. Sleep 2023.

14. Bilgel M, An Y, Walker KA, Moghekar AR, Ashton NJ, Kac PR et al. Longitudinal changes in Alzheimer’s-related plasma biomarkers and brain amyloid. Alzheimers Dement 2023.

15. Tang Y, Han L, Li S, Hu T, Xu Z, Fan Y, et al. Plasma GFAP in Parkinson’s disease with cognitive impairment and its potential to predict conversion to dementia. NPJ Parkinsons Dis 2023; 9(1): 23.

16. Bateman RJ, Wen G, Morris JC, Holtzman DM. Fluctuations of CSF amyloid-beta levels: implications for a diagnostic and therapeutic biomarker. Neurology 2007; 68(9): 666–669.

17. Dobrowolska JA, Kasten T, Huang Y, Benzinger TL, Sigurdson W, Ovod V et al. Diurnal patterns of soluble amyloid precursor protein metabolites in the human central nervous system. PLoS One 2014; 9(3): e89998.

18. Lucey BP, Fagan AM, Holtzman DM, Morris JC, Bateman RJ. Diurnal oscillation of CSF Abeta and other AD biomarkers. Mol Neurodegener 2017; 12(1): 36.

19. Palmqvist S, Janelidze S, Quiroz YT, Zetterberg H, Lopera F, Stomrud E et al. Discriminative Accuracy of Plasma Phospho-tau217 for Alzheimer Disease vs Other Neurodegenerative Disorders. JAMA 2020; 324(8): 772–781.

20. Mattsson-Carlgren N, Janelidze S, Palmqvist S, Cullen N, Svenningsson AL, Strandberg O et al. Longitudinal plasma p-tau217 is increased in early stages of Alzheimer’s disease. Brain 2020; 143(11): 3234–3241.

21. Huber H, Ashton NJ, Schieren A, Montoliu-Gaya L, Molfetta GD, Brum WS et al. Levels of Alzheimer’s disease blood biomarkers are altered after food intake-A pilot intervention study in healthy adults. Alzheimers Dement 2023.

