## Supplemental Tables for "P-tau217 and other blood biomarkers of dementia: variation with time of day"

### Supplementary tables

**Supplementary Table 1.** Mixed model results including age, sex, BMI, PSQI, and AHI in the model for the nine-time points.

| | Main effect (N = 21) | | | | | | | | | | | | | | | |
|---|---|---|---|---|---|---|---|---|---|---|---|---|---|---|---|---|
| Variable | Age | | Sex | | BMI | | PSQI | | AHI | | Group | | Time | | Group*Time | |
| | *F (DF)* | *p* | *F (DF)* | *p* | *F (DF)* | *p* | *F (DF)* | *p* | *F (DF)* | *p* | *F (DF)* | *p* | *F (DF)* | *p* | *F (DF)* | *p* |
| P-tau217 | 0.23 (1,12.9) | 0.640 | 2.01 (1,12.9) | 0.180 | 0.23 (1,12.9) | 0.639 | 0.77 (1,13) | 0.396 | 0.13 (1,12.9) | 0.722 | 6.06 (2,13) | **0.014** | 4.28 (8,21) | **0.0001** | 1.28 (16,121) | 0.219 |
| Aβ40 | 1.77 (1,12.9) | 0.206 | 0.04 (1,12.9) | 0.841 | 1.12 (1,12.9) | 0.310 | 0.03 (1,12.9) | 0.856 | 1.94 (1,12.9) | 0.187 | 0.1 (2,12.9) | 0.903 | 4.8 (8,121) | **<.0001** | 0.92 (16,121) | 0.553 |
| Aβ42 | 1.48 (1,12.9) | 0.246 | 0.16 (1,12.9) | 0.694 | 2.02 (1,12.9) | 0.179 | 0.99 (1,12.9) | 0.339 | 1.13 (1,12.9) | 0.308 | 0.51 (2,12.9) | 0.614 | 6.41 (8,121) | **<.0001** | 0.66 (16,121) | 0.830 |
| Aβ42/Aβ40 | 0.22 (1,13) | 0.644 | 0.25 (1,13) | 0.626 | 0.14 (1,13) | 0.718 | 1.38 (1,13) | 0.261 | 0.64 (1,13) | 0.438 | 0.79 (2,13) | 0.473 | 3.01 (8,21) | **0.004** | 0.99 (16,121) | 0.470 |
| GFAP | 5.45 (1,12.9) | **0.036** | 0.95 (1,12.9) | 0.349 | 0.04 (1,12.9) | 0.841 | 0.19 (1,12.9) | 0.669 | 0.48 (1,12.9) | 0.499 | 1.04 (2,12.9) | 0.383 | 1.88 (8,21) | 0.069 | 1.25 (16,121) | 0.242 |
| NfL | 3.94 (1,13) | 0.069 | 4.3 (1,13) | 0.059 | 0.18 (1,13) | 0.677 | 0.31 (1,13) | 0.586 | 0.61 (1,13) | 0.449 | 0.31 (2,13) | 0.742 | 2.05 (8,21) | **0.046** | 0.91 (16,121) | 0.557 |

**Supplementary Table 2.** Mixed model results including age, sex, BMI, PSQI, and AHI in the model for evening and morning time points.

| | | Main effect | | | | | | | | | | | | | | | |
|---|---|---|---|---|---|---|---|---|---|---|---|---|---|---|---|---|---|
| Variable | N | Age | | Sex | | BMI | | PSQI | | AHI | | Group | | Time | | Group*Time | |
| | | *F (DF)* | *p* | *F (DF)* | *p* | *F (DF)* | *p* | *F (DF)* | *p* | *F (DF)* | *p* | *F (DF)* | *p* | *F (DF)* | *p* | *F (DF)* | *p* |
| P-tau217 | 21 | 0.22 (1,12.9) | 0.647 | 2.38 (1,13) | 0.147 | 0.1 (1,12.9) | 0.757 | 0.76 (1,12.9) | 0.399 | 0.05 (1,12.9) | 0.819 | 5.23 (2,12.9) | **0.022** | 7.84 (1,17) | **0.0123** | 3.7 (2,17) | **0.046** |
| Aβ40 | 38 | 0.9 (1,29) | 0.350 | 0.11 (1,29.3) | 0.744 | 0.75 (1,29) | 0.393 | 0.02 (1,29) | 0.883 | 0.07 (1,29) | 0.795 | 0.66 (2,29.2) | 0.523 | 3.91 (1,32.7) | 0.0566 | 0.43 (2,32.7) | 0.657 |
| Aβ42 | 38 | 0.06 (1,29.4) | 0.804 | 0.04 (1,29.6) | 0.843 | 1.96 (1,29.4) | 0.172 | 0.26 (1,29.4) | 0.614 | 0 (1,29.4) | 0.963 | 0.47 (2,29.6) | 0.632 | 1.87 (1,32.8) | 0.1806 | 0.54 (2,32.8) | 0.589 |
| Aβ42/Aβ40 | 38 | 0.56 (1,29.8) | 0.459 | 0.04 (1,30) | 0.836 | 0.67 (1,29.8) | 0.420 | 0.88 (1,29.8) | 0.356 | 0.06 (1,29.8) | 0.807 | 0.07 (2,29.9) | 0.937 | 1.4 (1,33.1) | 0.245 | 0.17 (2,33.1) | 0.843 |
| GFAP | 38 | 8.73 (1,29.4) | **0.006** | 0 (1,29.4) | 0.957 | 0.01 (1,29.4) | 0.921 | 0.08 (1,29.4) | 0.784 | 0.03 (1,29.4) | 0.862 | 0.62 (1,29.4) | 0.546 | 2.78 (1,29.4) | 0.105 | 2.75 (1,29.4) | 0.079 |
| NfL | 38 | 11.44 (1,29.9) | 0.002 | 1.34 (1,30) | 0.256 | 0.43 (1,29.9) | 0.516 | 0.16 (1,29.9) | 0.694 | 0.07 (1,30) | 0.800 | 0.71 (2,30) | 0.500 | 0.28 (1,33.3) | 0.599 | 0.71 (2,33.3) | 0.500 |
